# Computational methods for continuous eye-tracking perimetry based on spatio-temporal integration and a deep recurrent neural network

**DOI:** 10.1101/2020.05.22.20106401

**Authors:** Alessandro Grillini, Alex Hernández-García, Remco J. Renken, Giorgia Demaria, Frans W. Cornelissen

**Affiliations:** Department of Ophthalmology, University Medical Center Groningen, Hanzeplein 1 9713 GZ Groningen, The Netherlands; Institute of Cognitive Science, University of Osnabrück, 27 Wachsbleiche, 49090 Osnabrück, Germany; Cognitive Neuroscience Center, Department of Biomedical Sciences of Cells and Systems, University Medical Center Groningen, Hanzeplein 1 9713 GZ Groningen, The Netherlands

## Abstract

Perimetry, the mapping of the sensitivity of different visual field locations, is an essential procedure in ophthalmology. Unfortunately, standard automated perimetry (SAP), suffers from some practical issues: it can be tedious, requires manual feedback and a high level of patient compliance. These factors limit the effectiveness of perimetry in some clinical populations. In an attempt to remove some of these limitations, alternatives to SAP have been tried based on tracking eye movements. These new approaches have attempted to mimic SAP, thus presenting stimuli on a fixed grid, and replacing manual by ocular responses. While this solves some issues of SAP, these approaches hardly exploit the high spatial and temporal resolution facilitated by eye-tracking. In this study, we present two novel computational methods that do tap into this potential: (1) an analytic method based on the spatio-temporal integration of positional deviations by means of Threshold Free Cluster Enhancement (TFCE) and (2) a method based on training a recursive deep artificial neural network (RNN). We demonstrate that it is possible to reconstruct visual field maps based on continuous gaze-tracking data acquired in a relatively short amount of time.

## 1 Introduction

The assessment of the quality of the visual field (also called perimetry) is a staple of ophthalmic evaluation. The presence of a scotoma, a region of the visual field with reduced sensitivity, is a very characteristic symptom of diseases and disorders such as macular degeneration (Tolentino et al., 1994), glaucoma (Heijl & Bengtsson, 1996), hemianopia (Williams, 1997) and several forms of retinopathy (Greite et al., 1981; Voipio & Karjalainen, 2009; Alexander et al., 2004).

The current gold standard in the diagnostic assessment of the visual field is Standard Automated Perimetry (SAP) (Barton & Benatar, 2003). The main advantages of SAP are a thorough evaluation of multiple visual field locations, and relatively easy-to-interpret results that are normalized with respect to an age-matched population. However, the approach also has several limitations: the task is complicated for people with limited cognitive capabilities, demands patient compliance (Szatmáry, 2002), and requires maintaining a stable fixation for prolonged periods of time. Furthermore, patient performance is affected by learning (Schultz, 1990; Wild et al., 2006) and fatigue (Johnson et al., 1988), as well as the expertise of the operator (Montolio et al., 2012). Together, these constraints limit the effectiveness of SAP, particularly in clinical and rehabilitation contexts such as when dealing with children (Walters et al., 2012), the elderly and/or cognitively impaired patients (Gangeddula et al., 2017).

To overcome some of these issues, various groups have implemented variants of SAP in which eye-movements substituted the manual responses required on each trial of SAP (Jones et al., 2019; Pel et al., 2013). While this already simplifies the task, the resulting procedure is still prone to issues related to patient compliance and the requirement to stably fixate. Moreover, by retaining the trial- and grid-based approach of SAP, the full potential of the high spatial and temporal resolution facilitated by eye tracking is not exploited. For these reasons, we recently proposed a novel eye-movement-based approach that completely removes the trial-based aspect of SAP, in favor of a continuous assessment of oculomotor behavior over time (Grillini et al., 2018). This technique exploits the ocular following response, thus reducing the attention and compliance demanded from a patient. Simultaneously, the continuous assessment reduces test time considerably. In our study we showed that, based exclusively on the spatio-temporal properties of the eye-movements made during a short continuous tracking task, it is possible to classify a visual field defect (VFD). A limitation of our initial approach was that it could only classify a scotoma as belonging to one of the scotoma shape classes on which the machine classifier had been trained, and thus not reconstruct its actual location and shape. The absence of this type of information hinders a more general application of this technique in clinical and rehabilitation practice.

To overcome this limitation, here we propose two methods for analyzing continuous gaze data acquired during a tracking task that enables reconstructing the visual field including any VFD present.

Our first method is based on the intuition that, compared to a healthy participant, a patient with a VFD will make larger and more prolonged tracking errors if the stimulus falls within their scotomatous region. In essence, the method applies Threshold-Free Cluster Enhancement (TFCE) (Smith & Nichols, 2009) to perform a spatio-temporal integration of a time series of eye-stimulus positional deviations (the signal). This results in a weighted integration of the height and extent of the signal, which in our case represent space (positional deviation) and time (duration of the deviation until it is corrected), respectively. Next, we reconstruct the visual field and presence of any scotoma, by back-projecting the TFCE values into visual field space. We will refer to it as the TFCE method.

The second method is based on training a recurrent deep artificial neural network. It constitutes a data-driven approach that learns features from the time-series of gaze location data collected during the tracking task. This method makes no explicit hypotheses about the underlying relationship between eye movements and scotoma characteristics, and learns how the presence of a scotoma influences a participant’s visual behavior during the tracking task. The algorithm is a 7-layer recurrent neural network whose weights are optimized to minimize its time-point-wise predictions on a set of labeled training data (obtained on the basis of gaze-contingent simulations of scotomas in known locations). Once trained, the model is able to accurately predict whether the discrepancy between eye and stimulus positions, at a given time point, is caused by the presence of a scotoma. We reconstruct the visual field and presence of any scotoma by back-projecting the RNN predictions into visual field space. We will refer to it as the recurrent neural network (RNN) method.

Based on simulated gaze-contingent scotoma data previously reported on in Grillini et al. (2018), we show that both methods are able to reconstruct the shape of the scotomatous regions. While further improvements are still desirable, these methods constitute a stepping stone towards the design of an easy and effective eye-movement-based perimetry. This technique, having fewer limitations than SAP, can complement it in clinical and rehabilitation practices.

## 2 Methods

### 2.1 Data acquisition

Participants, apparatus, stimuli, and procedures of data acquisition have previously been described in Grillini et al. (2018). To summarize, the stimulus comprised a small (0.5 deg) dot, displayed at one of two possible contrast levels (5% and 50%), moving along a random walk path with or without random displacements in order to induce saccades (the smooth pursuit and saccadic pursuit conditions, respectively). The gaze of the participants was recorded with an infra-red light-based video eye tracker while they tracked the stimulus with their eyes. During the experiment, the participants (N=50) were subjected to different kinds of simulated gaze-contingent visual field defects: no loss, central loss, peripheral loss, and hemifield loss (see Figure 1). Each trial, lasting 20 seconds, was repeated 6 times for each condition (2 contrast levels × 2 pursuit modalities) for a total of 24 trials and a total test time of 480 seconds. In order to reliably assess the quality of VF reconstructed using the TFCE- and RNN-based methods, we carried out a 5-fold cross-evaluation. To do this, we split the data from the 50 participants into 5 sets, each containing the data from 40 participants for training and 10 participants for testing.

**Figure 1:**
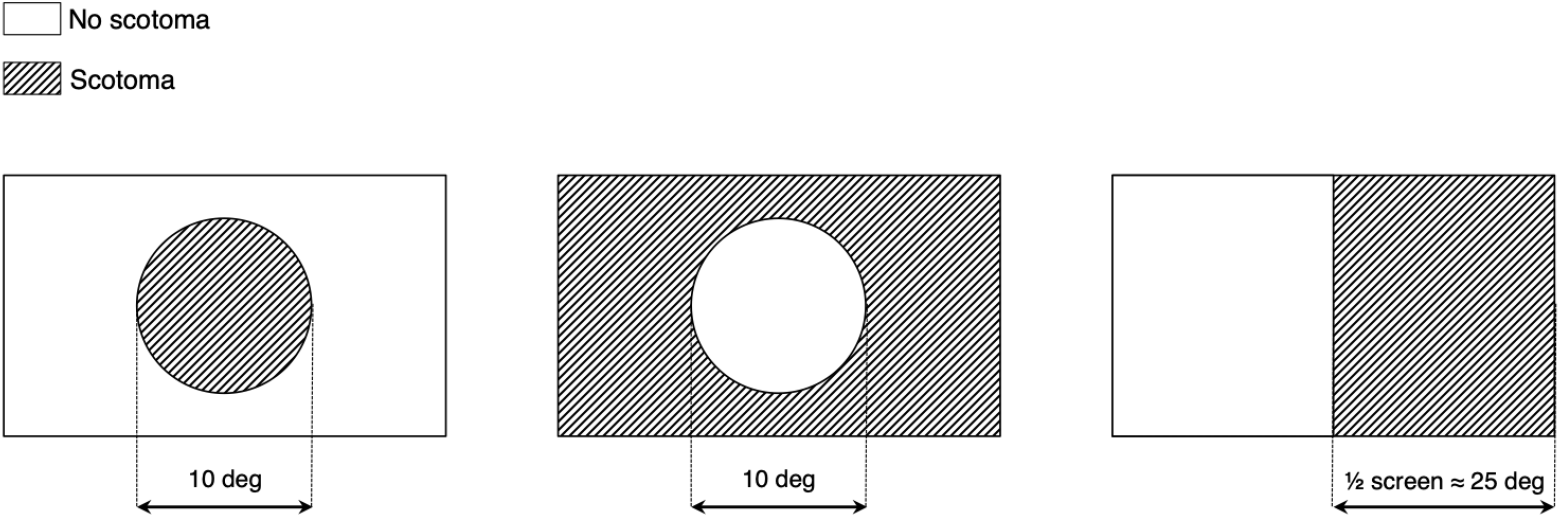
Examples of simulated visual field defects (VFD). From left to right: central loss, peripheral loss, hemifield loss. During the experiment, the VFD were applied in a gaze-contingent manner.

In order to assess the feasibility of these methods in a clinical setting, we further assessed three participants diagnosed with primary open-angle glaucoma (POAG) and three age-matched healthy control participants.

All participants gave their written informed consent prior to participation. The study was approved by the Medical Ethical Committee of the University Medical Center Groningen and the Ethics Committee of Psychology of the University of Groningen. The study followed the tenets of the Declaration of Helsinki.

### 2.2 Gaze-contingent simulated VFD

Details of the procedure to simulate VFD are described in Grillini et al. (2018). A schematic representation of the three types of simulated VFD used in this study is shown in Figure 1.

### 2.3 Method 1: Spatio-temporal integration of positional deviations by means of Threshold Free Cluster Enhancement (TFCE)

To simultaneously factor in the magnitude of the error and its duration, we applied to our data the Threshold Free Cluster Enhancement (TFCE) (Smith & Nichols, 2009). This algorithm, originally developed for the analysis of the hemodynamic response in functional neuroimaging, specifically helps to avoid the introduction of arbitrary thresholds when performing multiple-comparison corrections. In our context, we applied the algorithm to the time-series of positional deviations *D*, where each value is the Euclidean distance between the gaze location and the stimulus position at any given time point *t*. The positional deviations as a function of time are defined in Eq. 1:

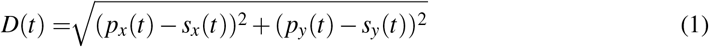

where *p*(*t*) and *s*(*t*) are the positions of the eye and the stimulus, respectively, divided into their horizontal (*x*) and vertical (*y*) components.

*D*(*t*) constitutes the input for the spatio-temporal integration performed with the TFCE equation, described in Eq. 2:

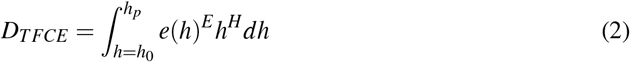

where e is the extent (temporal duration) and h is the height (spatial magnitude) of *D*(*t*) at a given point in time *t* (see Figure 2 for examples). This integral is implemented as a discrete sum using a finite step-size *dh*, (in our implementation *dh* = 1/2500*^th^* of the maximum of *D*(*t*)); *h*_0_ is the minimum of D(t) (which is always greater than or equal to 0), and *E* and *H* are the exponents. The resulting *D_TFCE_*(*t*) is a time-series of positional deviations weighted for their spatio-temporal integrated characteristics. Figure 2 shows some examples of using different *E* and *H* pairs: higher *E* values (red signals) enhance clusters with longer duration and suppress shorter ones; higher *H* values (blue signals) enhance the clusters with higher peaks and suppress the lower ones (Figure 2). We set these parameters to the recommended values of *E* = 2 and *H* = 0:5 (Smith & Nichols, 2009).

**Figure 2:**
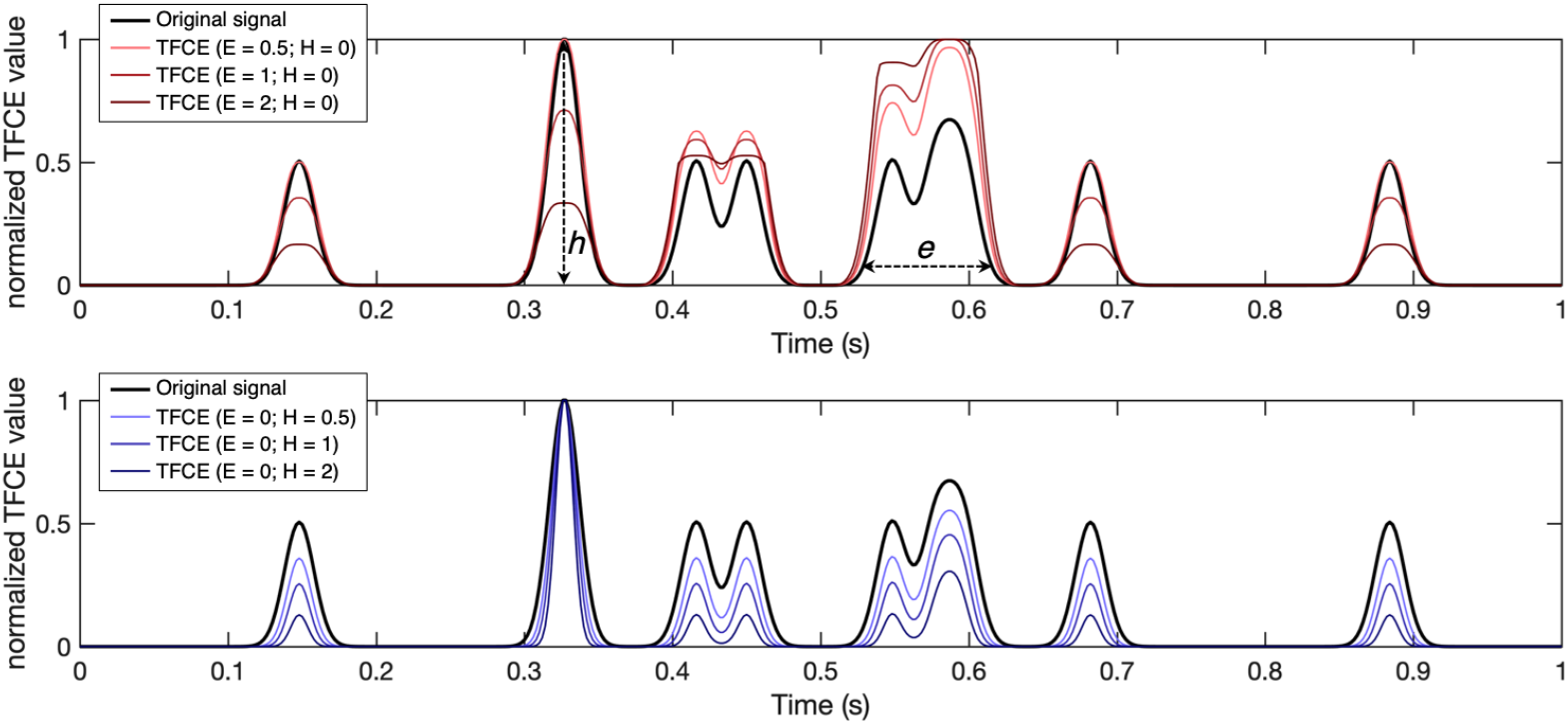
Examples of time series filtered using the TFCE algorithm with different combinations of parameters. A larger E weighs more the components of the original signal with longer temporal extent (*e*), defined as the time interval where the signal is consistently above a predefined minimum. Larger *H*, conversely, weighs more the highest components of the signal in terms of intensity (*h*). Note that the TFCE values here are normalized for visualization purposes only.

To label each value of *D_TFCE_*(*t*) as healthy or visual loss we apply the following algorithm.

First, in order to obtain the frequency distribution of all possible normative values (i.e. values of *D_TFCE_*(*t*) that should be considered healthy), *D_TFCE_*(*t*) is initially computed for every participant of the training dataset in the condition without simulated visual field loss, and the resulting values are aggregated with a histogram *F*.

Next, in order to choose the optimal boundary between “healthy” and “visual loss” values, we set a moving threshold λ*_n_*, such that *F*(λ*_n_*) = *n^th^* percentile of *F*. For each value of λ*_n_* we compute *B*(*t*;λ*_n_*) that is the binarized form of *D_TFCE_*(*t*) such as that *B*(*t*;λ*_n_*) = 0, if *D_TFCE_*(*t*) ≤ *F*(λ*_n_*) if “healthy” and *B*(*t*;λ*_n_*) = 1; if *D_TFCE_*(*t*) > *F*(λ*_n_*) if “visual loss”.

Finally, the visual field map is reconstructed as described in paragraph 2.5 “*Time-series back-projection in visual field space*”.

### 2.4 Method 2: Recursive Neural Networks (RNN)

In this method, we train a recursive neural network (RNN), as it is the most suitable known architecture to account for the temporal properties of the data (Rumelhart et al., 1986).

As training input **x**,, we use the time series of the eye gaze positions *p*(*t*) and the stimulus positions *s*(*t*), as well as the luminance contrast (low contrast = 0; high contrast = 1) and type of pursuit of the stimulus (smooth pursuit = 0; saccadic pursuit = 1).

As training output **y**, we use both the shape of the visual field defect (classified as *no loss, central loss, peripheral loss, and hemifield loss*) of the participants that generated each training sequence and, for each time time-point, whether the stimulus position lies in a location obstructed by the simulated scotoma.

As shown in Figure 3, the network consists of two streams that initially process the sequential data (*p*(*t*) and *s*(*t*)) and categorical data (high/low stimulus luminance contrast and smooth/saccadic pursuit) separately. In particular, the sequential stream contains three bidirectional recurrent Gated Recurrent Unit (GRU) layers (Cho et al., 2014) to effectively process the temporal dependencies of the sequential data. The outputs of both streams are then concatenated and used to jointly train two different softmax classifiers. One is trained to classify the shape of the visual field defect (no loss, central, peripheral, hemifield; analogously to what was reported by Grillini et al. (2018)), while the second one was trained to classify the visual field in a point-wise manner, i.e. “does the stimulus position in visual field space coordinates overlap with the scotoma?”.

**Figure 3:**
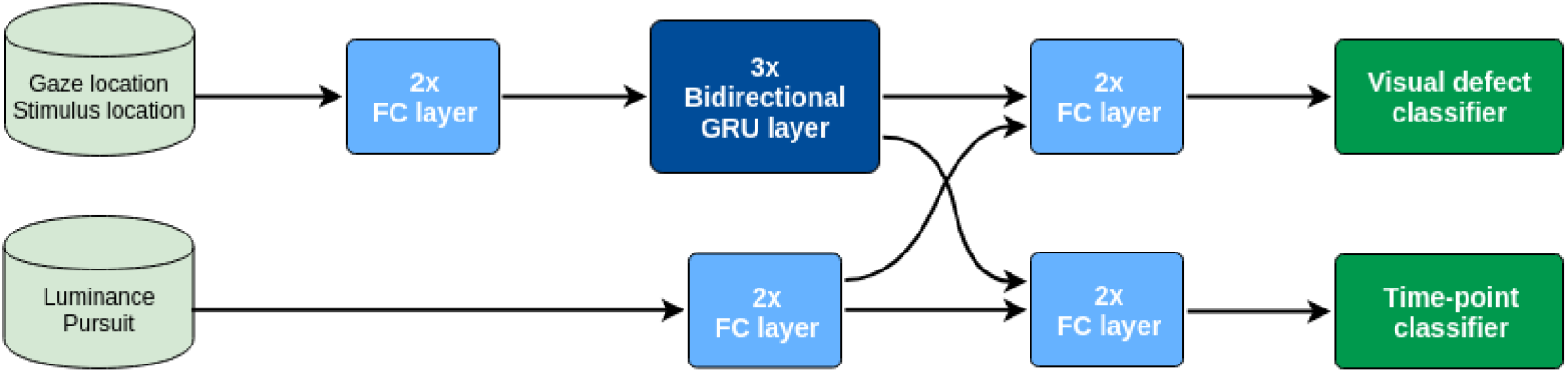
Schematic representation of the architecture of the deep recursive neural network.

We use the cross-entropy loss to define the cost function of the model, defined in Eq. 3:

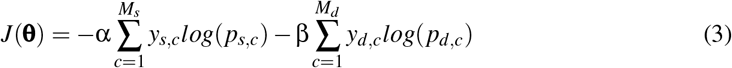

where *M* is the number of classes, *y* is the ground truth label and *p* is the predicted probability distribution, i.e. the output of each softmax classifier. Subscript *s* refers to the point-wise scotoma classifier and subscript *d* to the visual field defect shape classifier. In order to give priority to optimizing the reconstruction of the visual field, we set α = 0.75 and β = 0.25. The parameters 0 of the model are learned through mini-batch gradient descent, using RMSprop (Tieleman & Hinton, 2012), for 15,000 iterations with a batch size of *Bs* = 128.

The training batches are formed by first selecting *Bs* different sequences from the set of 20-seconds trials, originally sampled at 240 Hz. Then, we randomly sampled one sub-sequence of 4.17 seconds (1,000 time steps) from each sequence and finally down-sample them at 60 Hz (250 time steps). The stimulus contrast level and pursuit modality of the corresponding sequences are also added to the training batches.

This deep model can be regarded as a mapping **y** = *f* (**x**; θ), where

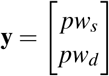, *pw_s_* being the point-wise scotoma prediction and *pw_d_* the visual field defect shape prediction of a sub-sequence **x**. In order to classify the visual field defect shape of one participant, we are interested here in *pw_d_*.

Since the data acquisition for one participant consists of 6 repetitions of 20-seconds trials for each luminance/contrast combination, we average the predicted output probability distributions of multiple sub-sequences. In particular, we average the predictions of the 6 × 2 × 2 = 24 down-sampled sequences. The predicted visual field defect for a participant s is thus defined by Eq. 4:

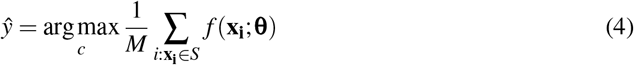

where *M* is the number of sub-sequences in the set of trials *S* of participant *s*.

### 2.5 Time-series back-projection into visual field space

Both the threshold-free cluster enhancement and the recurrent neural network outputs consist of binarized time-series *B*(*t*) where each entry has a value 0 if it is classified as not being obstructed by a scotoma and 1 if it is. Each entry also has associated a pair of *xy* coordinates, where x is the difference between horizontal gaze and stimulus positions at that time point and, analogously, *y* is the difference between vertical gaze and stimulus positions. These are retinotopic coordinates, meaning that they represent where the stimulus was with respect to the gaze of the participant. These coordinates are then binned into an *N* × *M* grid, where each square represents 1 deg^2^ of visual space and *N* and *M* are the dimensions of the visual field tested. Each square contains the percentage of occurrences that that specific location has been classified as being obstructed by a scotoma and gets color-coded accordingly. For visualization purposes, these retinotopic coordinates can be easily converted into polar coordinates (see Figure 6 and 7 for several examples).

A summary of the pipeline for visual field map reconstruction from gaze-tracking TFCE-filtered time-series is shown in Figure 4. The RNN visual field map reconstruction is analogous, with the binarized values of *B*(*t*) being provided by the outcome of the time-point classifier (Figure 3) rather than the threshold value *F*(λ*_n_*) applied to TFCE-filtered eye-tracking signals.

**Figure 4:**
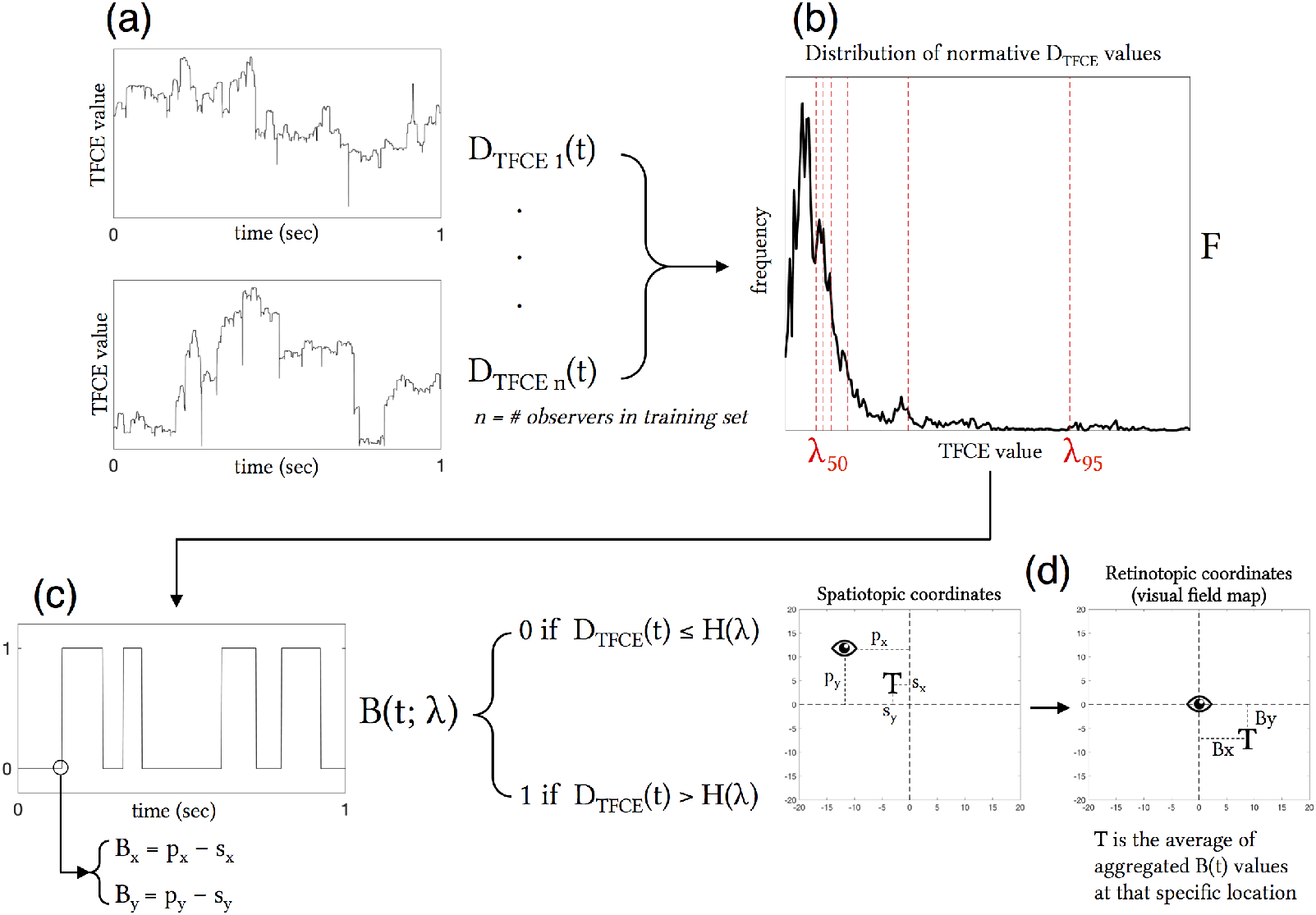
Schematic representation of the visual field map reconstruction algorithm pipeline. Starting from the TFCE-filtered time-series (**a**), a probability distribution *F* of all possible normative TFCE values is computed (**b**). For all percentiles λ*_n_* of the distribution *F*, a threshold value *F*(λ*_n_*) is defined and used to binarize the TFCE-filtered signal in to be analyzed (**c**). Each time point of the TFCE-filtered signal has associated the spatiotopic coordinates of gaze (*p_x_* and *p_y_*) and stimulus positions (*s_x_* and *s_y_*), which are converted into retinotopic coordinates *B_x_* and *B_y_*. The resulting mapped retinotopic coordinates (**d**) are associated with a target location *T* which contains the average of the aggregated *B*(*t*) values at that specific location, that is the expected probability that that specific location is affected by a scotoma. An analogous back-projection algorithm is implemented for the reconstruction of the visual field using the recurrent neural network, where the binarized time-series in (**c**) is defined by the output of the model instead of the threshold *F*(λ*_n_*).

Note that the back-projection into visual field space has been used to reconstruct the ground truth maps as well. In their case, the information about the presence or absence of the scotoma is known a priori, but the reconstruction is still necessary to ensure a proper spatial comparison between the ground truth and TFCE or RNN maps.

### 2.6 Evaluation of reconstructed maps accuracy and optimization of λ

In order to evaluate the accuracy of both the TFCE and the RNN methods, as well as to choose the optimal λ*_n_* parameter for the TFCE method, we compute a 2D Spearman rank correlation between the reconstructed visual field maps and their respective ground truth maps obtained with the known locations of the simulated scotomas.

The ground truth maps are obtained analogously as described in the Methods’ paragraph Time-series back-projection in retinotopic coordinates, using as an input the binarized time-series using the known location of the visual field loss instead of the predictions made by either TFCE or RNN.

First, we measure the correlation between ground truth and TFCE maps of the training set reconstructed using all possible values of λ*_n_* = [1,2,3,…, 100] (i.e. one for each percentile of the histogram *F*). Then the average between participants is computed for each simulated visual loss condition, followed by the grand average across conditions. The peak of the grand average corresponds to the optimal value of λ*_n_* that is used to reconstruct the maps of the test data. This procedure is repeated for each of the 5 folds of the cross-evaluation.

### 2.7 Clinical application of eye-tracking-based visual field reconstruction

In order to provide a proof-of-concept of the viability of these methods in a clinical setting, we asked 3 patients and 2 healthy controls participants with various degrees of visual field loss to perform the visual tracking task. We then compared the maps obtained with TFCE and RNN to those obtained with Standard Automated Perimetry (SAP) by means of the Humphrey Field Analyzer (HFA), using the SITA-Standard algorithm. The HFA was performed monocularly on the eye affected by primary open angle glaucoma (POAG) in the case of patients and in the dominant eye in the case of controls. We then compared the Mean Deviation (MD) as reported by the HFA (*MD_HFA_*) with an MD computed on the basis of the TFCE- and RNN-reconstructed maps (*MD_TFCE_* and *MD_RNN_*, respectively).

Since the stimulus used for the tracking task followed a random-walk path, we could not ensure a complete coverage of the whole visual field. Therefore, the computation of the *MD_TFCE_* and *MD_RNN_* values comprises a correction for visual field coverage. Our MD values are computed as follows:

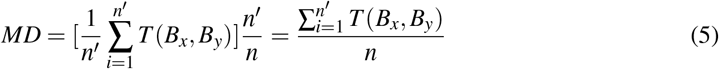

Where *n′* is the number of visual field locations sampled, *n* is the number of all possible visual field locations and *T* (*B_x_*, *B_y_*) is the expected probability that that specific location is affected by visual loss (see Figure 4).

## 3 Results

The results of the optimization of the TFCE parameter λ*_n_* based on the maximum average accuracy of each condition for each fold are shown in Figure 5, while Figure 6 shows the effect that adjusting λ*_n_* has on the visual field map reconstruction of a participant across all conditions.

**Figure 5:**
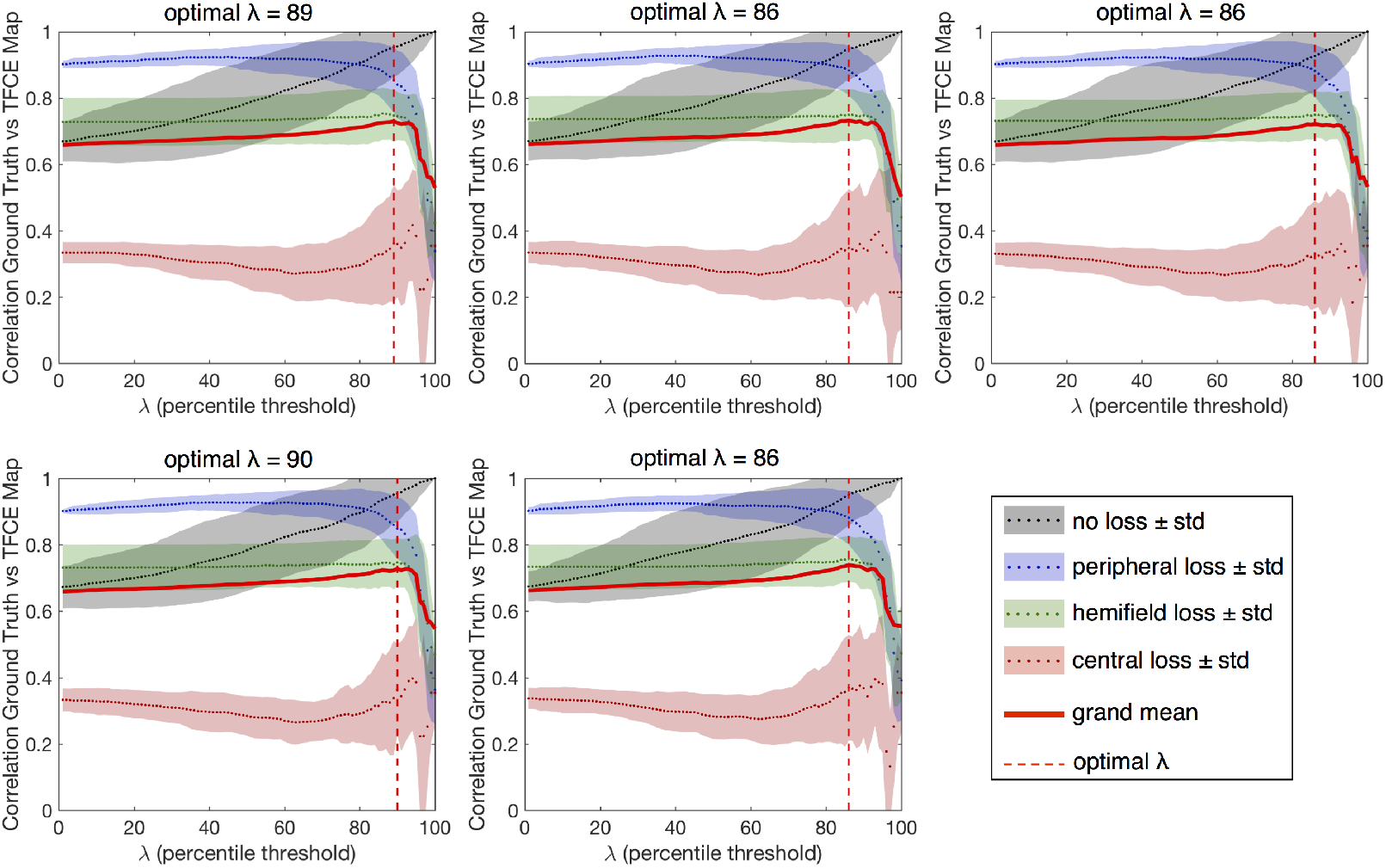
Results of the optimization of the λ parameter for each fold of the cross-evaluation. The optimal percentiles to be used as threshold between “healthy” and “impaired” TFCE values are in the range 86^th^ – 90^th^ (mean 87.4), corresponding to the peaks of the grand average between all tested conditions.

**Figure 6:**
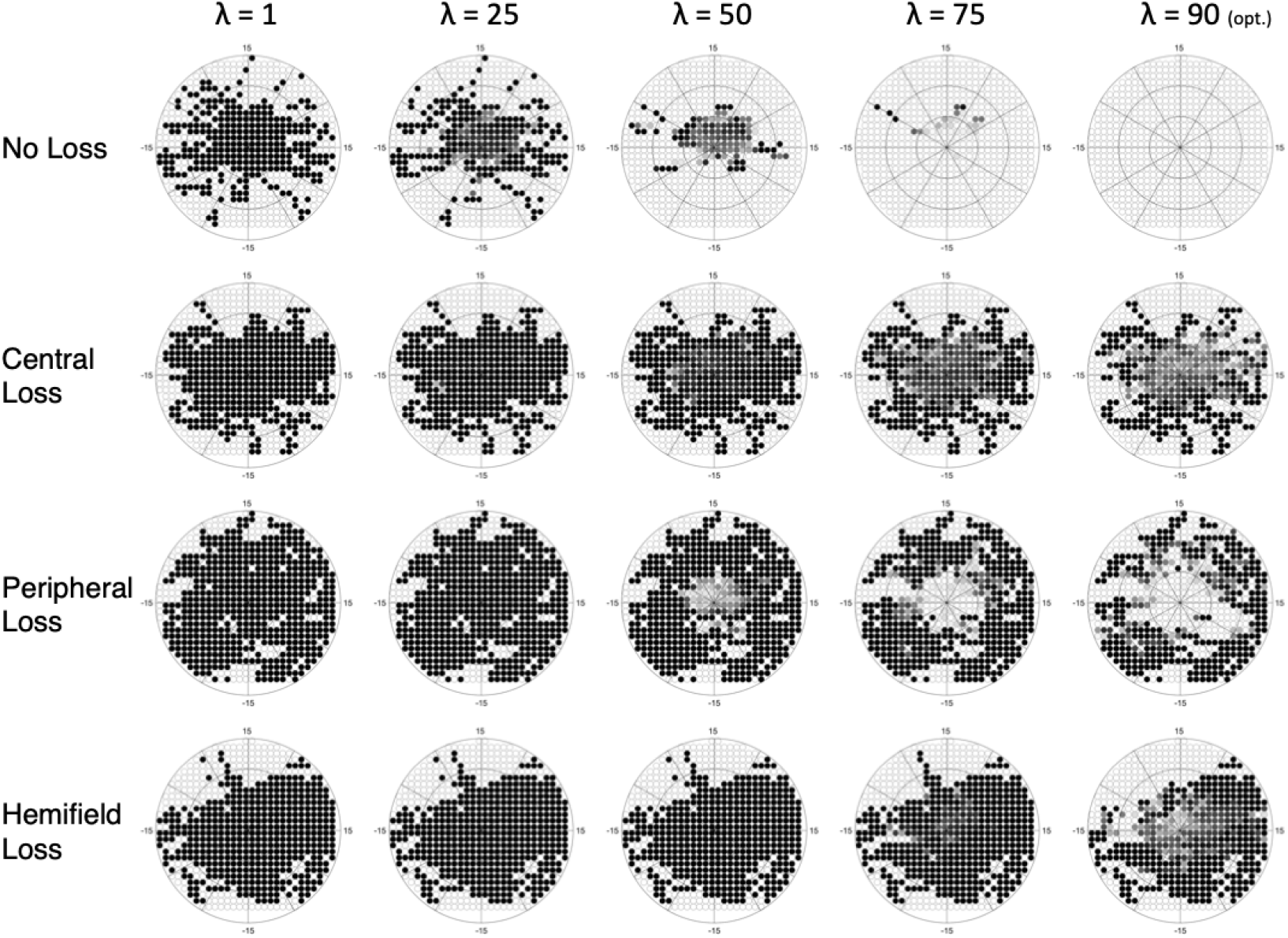
Example of the effect that adjusting the λ parameter has on a single participant, across all conditions. As the optimal λ is approached, the number of false positives in the “no loss” condition is minimized.

The results are very consistent across the 5 folds, with only the central loss condition showing minimal variability. The resulting optimal λ*_n_* is the average threshold determined for each of the five folds (#1: 89; #2: 86; #3: 86; #4: 90; #5: 86).

Next, in the test set, we reconstruct maps using both the TFCE (with the optimized λ*_n_*) and RNN methods. For each method separately we compute the 2D Spearman rank correlation with the respective ground truth reconstructed map. Figure 7 shows examples of reconstructed visual field maps for all simulated visual loss conditions applied to one random participant of the test set.

**Figure 7:**
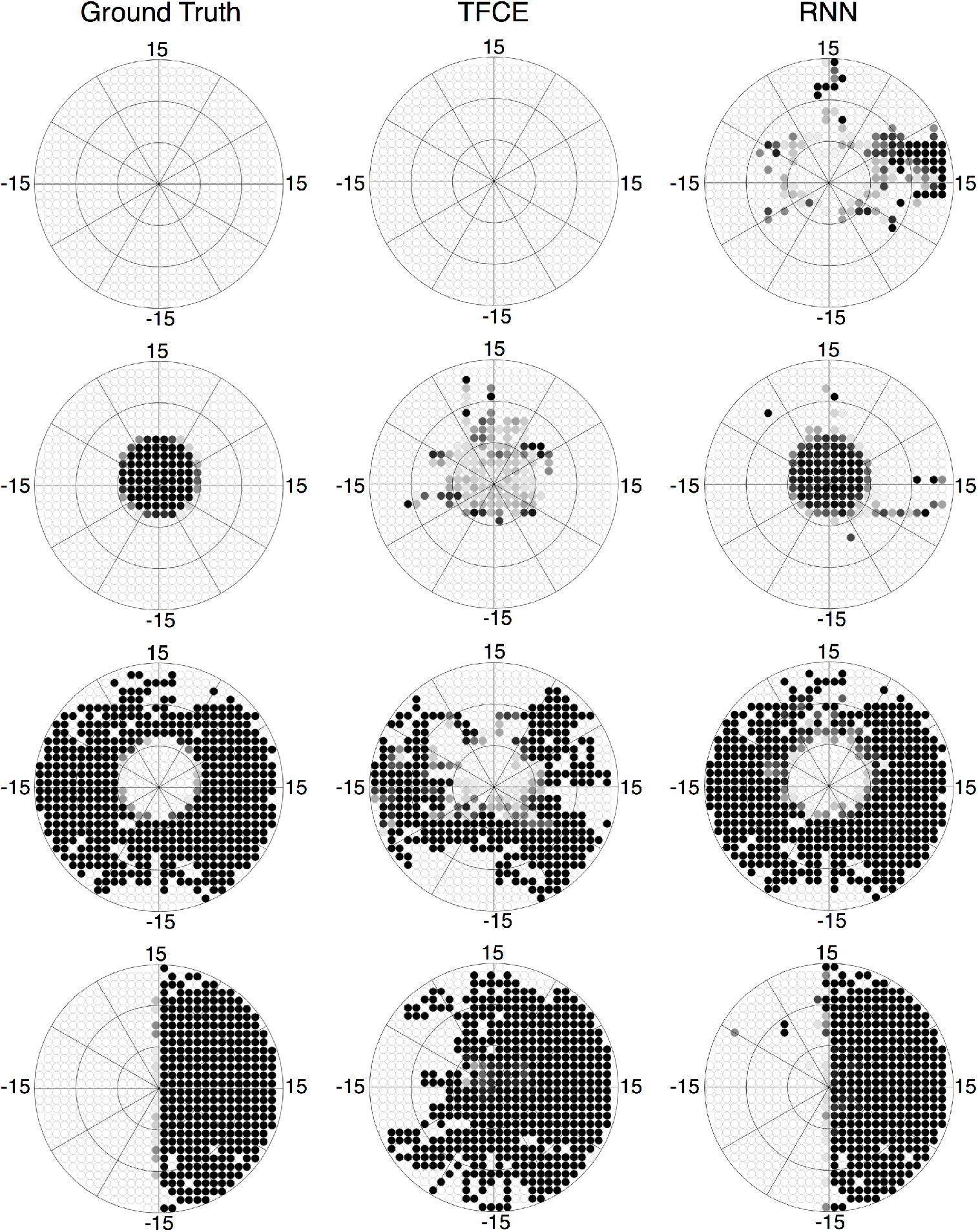
Reconstructed visual field maps for a single participant. Darker color indicates the higher probability of that point being affected by visual loss (white = 0%, black = 100%). The TFCE maps, although less accurate than the RNN ones, still allow to recognize the shape of the underlying scotomas. The RNN maps, on the other hand, closely resemble the ground truth maps.

We evaluated the overall performance of both the TFCE and RNN methods for all conditions tested, applying the 5-fold cross-evaluation for both methods using the same 40-10 split. Each fold uses its own optimised λ*_n_* as shown in Figure 5.

Figure 8 shows the performance for both the TFCE and RNN methods, quantified with the spatial distribution of False Positives and False Negatives (Figure 8-A), the False Positive Rate / False Negative Rate (Figure 8-B) and with their accuracy, computed as the 2D Spearman rank correlation between the reconstructed and ground truth maps (Figure 8-C).

**Figure 8:**
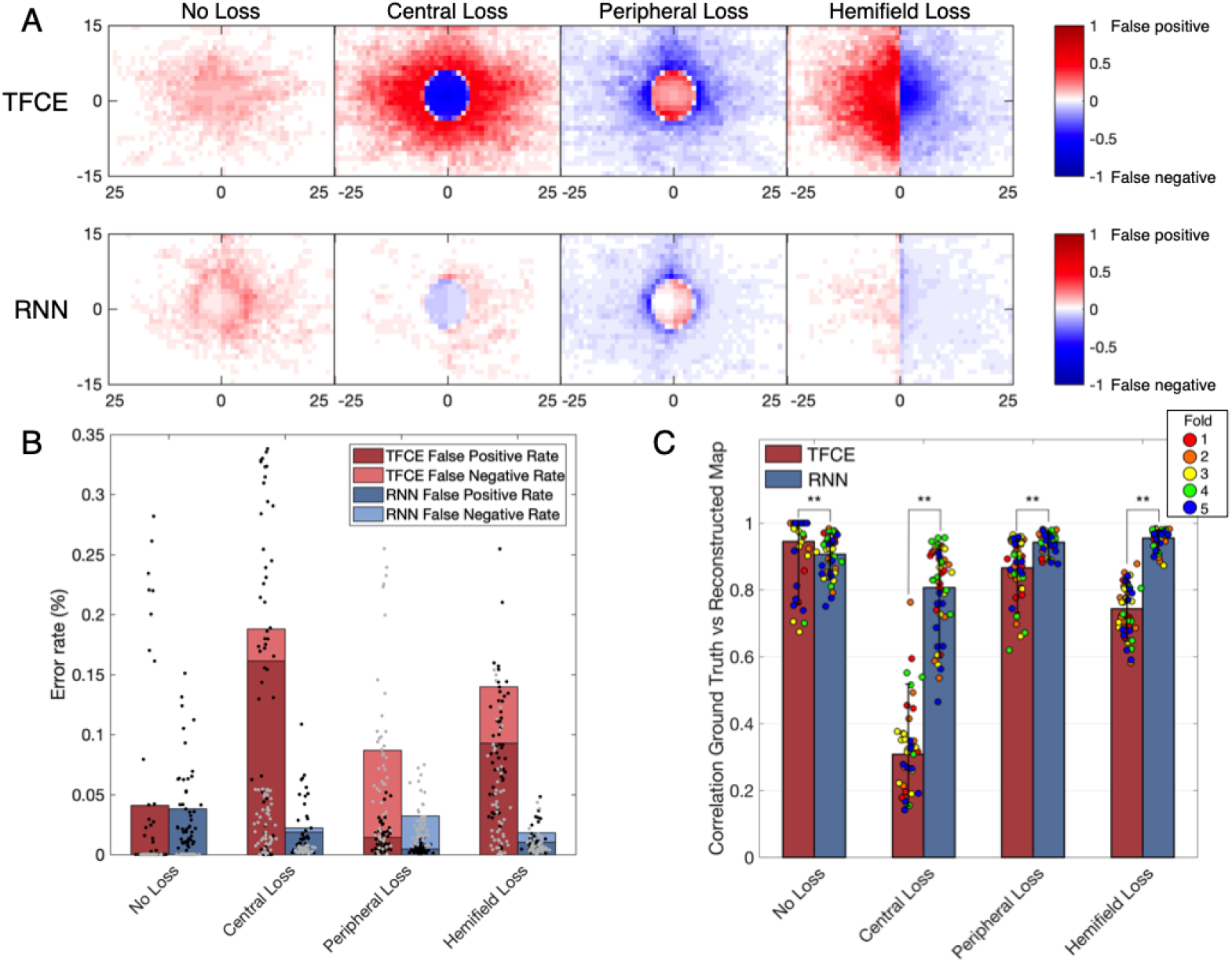
A. Spatial distributions of false positives and false negatives for each condition and for both TFCE and RNN methods. B. Total error rates combining false positive rate and false negative rate of TFCE and RNN. C. Comparison between TFCE and RNN visual field map reconstruction accuracies. The RNN method shows higher correlations with the ground truth as well as being more consistent than the TFCE method. Statistically significant differences (*p* < 0.01) are found across all tested conditions. Error bars show the 10*^th^* and 90*^th^* percentiles of each distribution.

A Kruskal-Wallis test (nonparametric one-way ANOVA) shows a statistically significant difference between the two methods for all observed conditions (all p-values < 0.01), where the RNN method proved to be more accurate than the TFCE in all but one condition (*No Loss*). Overall, the RNN method also showed less variability between participants (see Table 1)

**Table 1:** Accuracy and variability of the two methods TFCE and RNN.

|  | No Loss | Central Loss | Peripheral Loss | Hemifield Loss | Average |
| --- | --- | --- | --- | --- | --- |
| Accuracy TFCE | 0.9455 | 0.3122 | 0.8664 | 0.7435 | **0.7169** |
| Accuracy RNN | 0.9070 | 0.8066 | 0.9429 | 0.9555 | **0.9030** |
| IQR TFCE | 0.0693 | 0.1651 | 0.1038 | 0.1138 | **0.1141** |
| IQR RNN | 0.0693 | 0.1863 | 0.0303 | 0.0268 | **0.0782** |

Finally, to evaluate the generalizability of our approaches to a real clinical context, we compared the TFCE and RNN methods with the perimetry maps obtained with the Standard Automated Perimeter HFA, the current gold standard in perimetry. For this comparison, we tested participants with real visual field defects of different severity and compared the *MD_HFA_* to the *MD_TFCE_* and *MD_RNN_* computed as described in paragraph *”Application of eye-tracking visual field reconstruction to clinical data”* in Methods. For this comparison we took as optimal TFCE λ*_n_* the average between the optimal values of each fold. This λ*_n_* corresponds to the 87.4^th^ percentile of the distribution of all possible TFCE normative values. The clinical participants have been treated as a new independent test set, no re-training of the RNN model has been performed.

Contrary to the simulated scotomas, the TFCE method outperformed the RNN in this case, showing a significant correlation between its MD and the MD of the HFA. (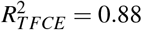, *p_TFCE_* = 0.0117; 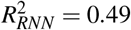, *p_RNN_* = 0.112).

## 4 Discussion

Our main conclusion is that it is possible to reconstruct visual field maps, including the location of a scotoma, based on eye-tracking data acquired with a method of continuous gaze tracking. We consider this a breakthrough proof-of-principle, as it indicates a pathway towards the design of a high-resolution, patient-friendly way to perform perimetry. Below, we discuss the merits (and limitations) of the two methods for visual field map reconstruction that we presented and tested in this study: (1) spatio-temporal integration of positional deviations performed with Threshold Free Cluster Enhancement (TFCE) and (2) recursive deep artificial neural network (RNN). Moreover, we will compare our techniques to other proposed methods for eye-tracking-based perimetry and discuss possible further improvements.

### 4.1 Continuous gaze-tracking allows the reconstruction of visual field maps

#### 4.1.1 TFCE

The spatio-temporal integration of positional deviations via Threshold Free Cluster Enhancement allows the reconstruction of visual field maps without requiring any prior knowledge about visual field defects. It is a method easy to implement, computationally inexpensive, and biologically-plausible: a loss of sensitivity in the visual field is associated with lower accuracy and higher delays of the eye-movements landing in the impaired region. This is consistent with findings from previous studies involving patients with central (Van der Stigchel et al., 2013) and peripheral (Burton et al., 2014) visual field defects.

The TFCE method performed quite well in reconstructing visual field maps with no loss or with peripheral loss (accuracies of 0.95 and 0.87, respectively), while it fared less optimally in reconstructing central losses and hemifield losses (accuracies of 0.31 and 0.74, respectively). A plausible reason for this discrepancy is the heavy foveal bias that is inherent to continuous tracking tasks: to accurately track the stimulus the observer must keep it as close as possible to the centermost part of their visual field. In case of occlusion of the fovea, this is not possible, leading to prolonged errors that never allow the positional deviations to return to zero (i.e. when the eye is on the target). In the TFCE algorithm, this results in an erroneous definition of the baseline (the *h*_0_ parameter in Eq. 2) which in turn leads to an increased error rate towards the center of the visual field for central loss and hemifield loss conditions (Figure 8-A and 8-B).

Furthermore, although this method fares well in detecting a scotoma in its expected location, it struggles in precisely defining the edges of the defect (for examples see Figure 7, second and fourth rows). This can be due to the presence of compensatory eye movement strategies in the observer or the use of a different preferred retinal locus in the presence of visual field defects, whether real (Coeckelbergh et al., 2002) or artificial (Cornelissen et al., 2005; McIlreavy et al., 2012).

Despite these limitations, the maps reconstructed with this method corresponded well to those reconstructed using SAP (Figure 9) and their respective MD values significantly correlated with each other, showing promising potential for the generalizability into clinical use.

**Figure 9:**
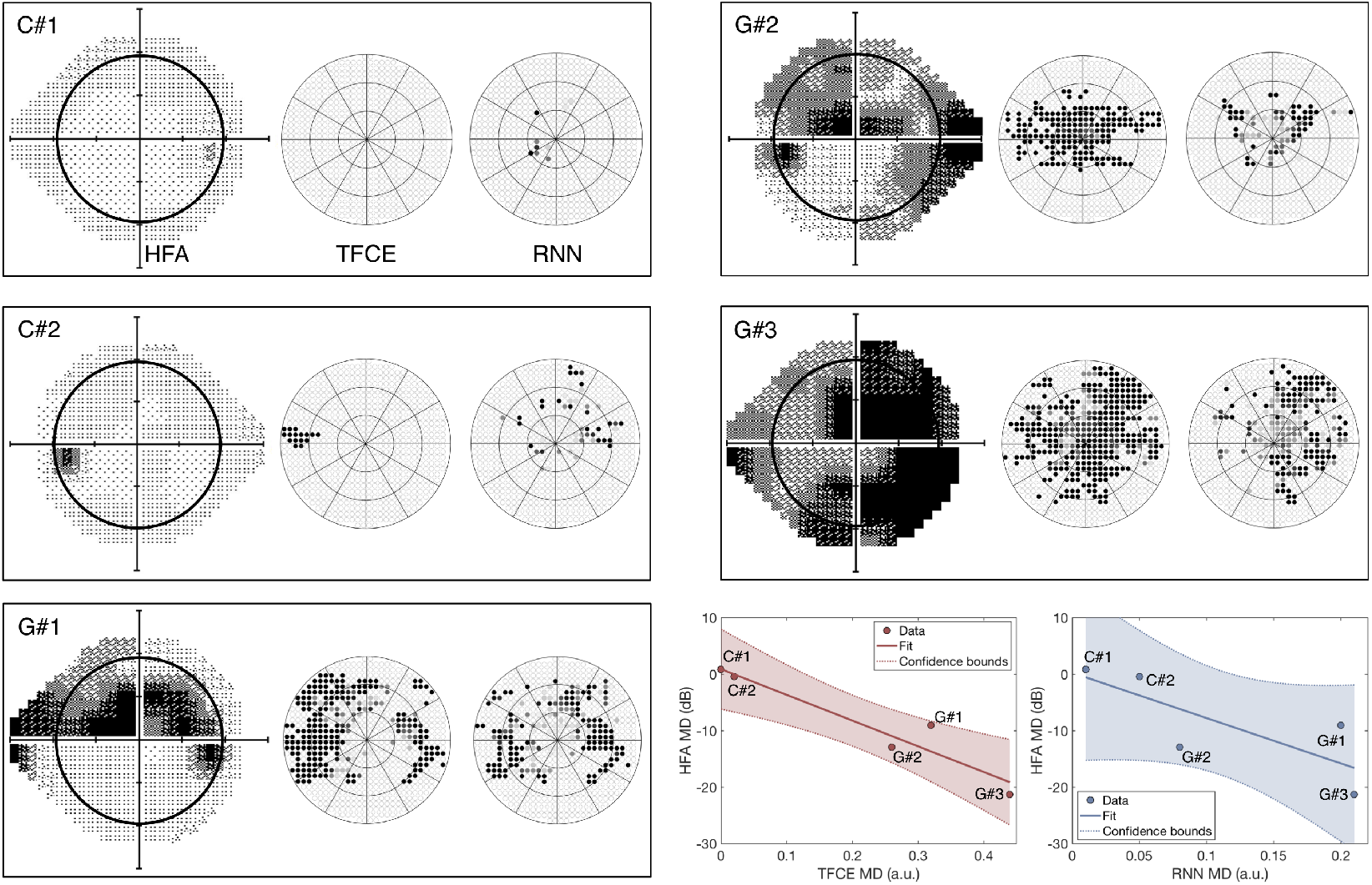
Perimetric maps of the five patients tested with our continuous tracking test. The TFCE maps are reconstructed using as λ*_n_* the average of the values obtained by each individual fold (see Figure 5). C#1 and C#2 are healthy controls, while G#1, G#2, and G#3 and patients previously diagnosed with Primary Open Angle Glaucoma. The black circle within the HFA maps represents the portion of the visual field covered by the TFCE and RNN maps. The TFCE method shows a significant correlation between its Mean Deviation index and the one obtained with the HFA, whereas the RNN method does not (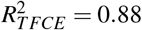, *p_TFCE_* = 0.0117; 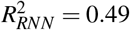, *p_RNN_* = 0.112).

#### 4.1.2 RNN

The reconstruction of visual field maps by means of the RNN method proved to be highly accurate with simulated scotomas, with an average accuracy across conditions above 0.90. This method, however, did not show significant correlations with the maps of POAG patients obtained using SAP. This could be due to the way the network was trained.

Using a limited number of predefined scotoma shapes made the RNN optimal at reconstructing similar scotomas (with an average accuracy above 0.90), but not as effective in dealing with new shapes not encountered before. This is also evident from the spatial distributions of false positives and false negatives (Figure 8-A, lower panel), where the RNN reveals having a clear internal representation of the four scotoma shapes used in the training data.

Another aspect that limits the implementation of the RNN in a clinical setting is that, in its current form, it requires training data for which the location of the scotoma is known. This is easily achievable using simulated gaze-contingent scotomas, but with actual patients, it is impossible to establish their objective ground truth. While comparison to SAP is an obvious approach as it is the current gold standard, this method can be unreliable when testing moderate to severe visual loss (Gardiner et al., 2014), cognitively impaired (Diniz-Filho et al., 2017), or very young patients (< 6 yrs old) (Patel et al., 2015; Tschopp et al., 1998), leading to a distorted ground truth that would not constitute good training data.

We must note though that most of these limitations can potentially be overcome by re-designing the way the training data is acquired. In the section 4.3 “*Current limitations and future improvements*”, we propose possible solutions to the training issue and other problems.

### 4.2 Comparison with existing tools for eye-tracking-based perimetry

The rationale behind the development of a perimetric tool based on continuous gaze tracking is rooted in the previous evidence that kinetic perimeters, i.e. devices such as the Goldmann Perimeter and the Octopus 900 where the probing stimulus is moving, outperform SAP both in reliability and ease of use in patients aged 5 to 8 (Patel et al., 2015). This suggests that the use of moving stimuli in perimetry, although considered not optimal for the general population, might be relevant within specific clinical contexts, and eye-tracking techniques constitute a fertile ground to explore this possibility.

We are not the first to propose eye tracking as a means towards removing some critical aspects of SAP, such as its high cognitive load and the need for manual feedback (Kim et al., 1995; Jones et al., 2019; Pel et al., 2013). In fact, even the counting of fixation losses and determining blink frequency can be seen as an elementary form of eye-tracking that is used to improve the reliability of SAP (Ishiyama et al., 2015; Asaoka et al., 2019).

So far, all existing tools for eye-tracking-based perimetry employ the same working principles of conventional perimetry with the primary difference being using ocular responses instead of manual ones. The patient is still asked to repeatedly answer the question “do you see the stimulus?”, and the answer is provided by the landing (or not) of a saccade within a Region-of-interest (ROI) around the target (Kim et al., 1995; Jones et al., 2019), or by the saccadic latency to fall within the ROI of a displaced target (Pel et al., 2013).

This approach comes with the advantage of allowing a precise sensitivity threshold estimation for each tested visual field location but has two major downsides. First, since each point needs to be tested individually and repeatedly, the spatial resolution is intimately interrelated to the available testing time. Second, discrete eye movement perimetry is heavily reliant on optimal instrument calibration: if the average calibration error exceeds the ROI radius of each target, it is very well possible to have completely invalid maps.

Our method based on continuous tracking is designed to be less affected by these issues. First, as each time sample contributes to the final map and spatial binning is applied only *a posteriori*, even without any smoothing, it is possible to obtain detailed maps (see Figures 6, 7, and 9).

Second, although still partially affected by calibration issues, our gaze-based method of visual field mapping can still allows for accurate classification of visual field defects. Our method is based on the same stimuli as used in a previous study by Grillini et al. (2018), where spatio-temporal features were used to classify the shape of the underlying visual field defect. The categorical classification in that study was performed by training a simple decision tree with the features explicitly extracted from the gaze and stimulus data. The temporal features are affected minimally by poor calibration and still yield sufficient information about the type of scotoma.

In the present study, these features are not made explicit, but their categorical classification is still performed by our neural network (see Figure 3, upper stream). As the temporal dependencies are taken into account by the long short-term memory properties of the bidirectional GRUs layers (Cho et al., 2014), the network should be able to perform a satisfactory visual field defect classification (not reconstruction) also in the presence of rather poor calibrations. Future studies should investigate this possibility.

### 4.3 Current limitations and future improvements

While we believe that the results presented constitute a promising proof-of-concept of the viability of continuous-gaze-tracking perimetry, their implementation into a clinical setting would still require a number of improvements, both in the acquisition and analysis of gaze data.

First, we trained the recurrent neural network using simulated visual field defects in order to establish a ground truth for the presence vs. absence of a scotoma. While this is not feasible with real scotomas, an alternative is to train the RNN with more realistic and diverse visual loss simulations. The ones that we used were either masking the stimulus completely or not at all, as well as having stereotyped shapes lacking all the idiosyncrasies that can be present in actual visual field impairments. Training the RNN with different (more organic) shapes with multiple levels of contrast reduction could provide a significant improvement over our present results.

Second, the TFCE method requires two parameters to be defined beforehand: H and E, representing the weights to be attributed to the height and to the extent of the positional deviations, respectively. These parameters are typically chosen empirically, and in the present study, we adopted the values recommended by the original authors Smith and Nichols, 2009 (Smith & Nichols, 2009). A future implementation of this method can comprise a preliminary optimization phase in which the parameters H and E are chosen analytically using machine learning. Possible computational approaches for the optimization of the TFCE hyperparameters would be grid-search cross-validation (Abu-Mostafa et al., 2012) or Bayesian optimization (Brochu et al., 2010).

Third, the optimization of the stimulus properties. The current stimulus is the same used to extract the spatio-temporal properties of eye movements for neuro-ophthalmic screening (Grillini et al., 2020), but it can be further optimized to perform visual field assessment. For instance, properties such as luminance contrast, speed, and trajectory of the moving target can be adjusted with the goal of maximizing visual field coverage and minimizing central bias.

Fourth, the simulated visual loss data used for training the RNN (healthy controls) and the clinical data (glaucoma patients) used to test it were acquired from two groups with different age ranges (20-30 vs 70-80). The different age ranges may have had an effect on eye movements even in the absence of a clinical condition (Rottach et al., 1996).

Lastly, an additional improvement could be to take into account the physiology of the retina and model H and E according to the “hill of vision” (Jacobs & Patterson, 1985). In this case, different values of H and E could be determined for sections of the retina at different eccentricity, such as the central peak (0-10 degrees), the mid-plateau (15-25 degrees) and the peripheral decay (above 25 degrees) and combined with the hyperparameter optimization methods mentioned above for best results.

### 4.4 Conclusions

We developed and proposed two methods that enable the reconstruction of visual field maps by estimating retinal sensitivity using continuous gaze-tracking data: (1) spatio-temporal integration of positional deviations performed with Threshold Free Cluster Enhancement (TFCE) and (2) recursive deep artificial neural network (RNN). The two methods possess complementary qualities (and downsides): the TFCE is biologically-plausible and computationally efficient while the RNN is remarkably accurate when provided with proper training data. We conclude that both methods can contribute to making gaze-based perimetry more viable in the future.

## Data Availability

Data is available upon request to the corresponding author.

## 4.5 Acknowledgments

This project has received funding from the European Union’s Horizon 2020 research and innovation programme under the Marie Sklodowska-Curie grant agreements No 641805 (“NextGenVis”) and No 675033 (“EGRET”). We are thankful to Nomdo Jansonius for his valuable advice and support.

## References

Abu-Mostafa, Y. S., Magdon-Ismail, M., and Lin, H.-T. Learning from Data: A Short Course. AML Books, January 2012.

Alexander, K. R., Barnes, C. S., Fishman, G. A., Pokorny, J., and Smith, V. C. Contrast-Processing deficits in Melanoma-Associated retinopathy. Investigative Opthalmology & Visual Science, 45 (1):305, 2004.

Asaoka, R., Fujino, Y., Aoki, S., Matsuura, M., and Murata, H. Estimating the reliability of glaucomatous visual field for the accurate assessment of progression using the Gaze-Tracking and reliability indices. Ophthalmology Glaucoma, 2(2):111–119, 2019.

Barton, J. J. S., and Benatar, M. Automated perimetry (humphrey field analyzer). Field of Vision, pp. 45–69, 2003.

Brochu, E., Cora, V. M., and de Freitas, N. A tutorial on bayesian optimization of expensive cost functions, with application to active user modeling and hierarchical reinforcement learning. CoRR, abs/1012.2599, 2010. URL http://arxiv.org/abs/1012.2599.

Burton, R., Smith, N. D., and Crabb, D. P. Eye movements and reading in glaucoma: observations on patients with advanced visual field loss. Graefes Arch. Clin. Exp. Ophthalmol., 252(10):1621–1630, October 2014.

Cho, K., van Merrienboer, B., Gulcehre, C., Bahdanau, D., Bougares, F., Schwenk, H., and Bengio, Y. Learning phrase representations using RNN Encoder-Decoder for statistical machine translation. Proceedings of the 2014 Conference on Empirical Methods in Natural Language Processing (EMNLP), 2014.

Coeckelbergh, T. R. M., Cornelissen, F. W., Brouwer, W. H., and Kooijman, A. C. The effect of visual field defects on eye movements and practical fitness to drive. Vision Res., 42(5):669–677, March 2002.

Cornelissen, F. W., Bruin, K. J., and Kooijman, A. C. The influence of artificial scotomas on eye movements during visual search. Optom. Vis. Sci., 82(1):27–35, January 2005.

Diniz-Filho, A., Delano-Wood, L., Daga, F. B., Cronemberger, S., and Medeiros, F. A. Association between neurocognitive decline and visual field variability in glaucoma. JAMA Ophthalmol., 135 (7):734–739, July 2017.

Gangeddula, V., Ranchet, M., Akinwuntan, A. E., Bollinger, K., and Devos, H. Effect of cognitive demand on functional visual field performance in senior drivers with glaucoma. Front. Aging Neurosci., 9:286, August 2017.

Gardiner, S. K., Swanson, W. H., Goren, D., Mansberger, S. L., and Demirel, S. Assessment of the reliability of standard automated perimetry in regions of glaucomatous damage. Ophthalmology, 121(7):1359–1369, July 2014.

Greite, J.-H. H. Greite, J. P. Zumbansen, H., and Adamczyk, R. Visual field in diabetic retinopathy (DR). Fourth International Visual Field Symposium Bristol, April 13–16,1980, pp. 25-32, 1981.

Grillini, A., Ombelet, D., Soans, R. S., and Cornelissen, F. W. Towards using the spatio-temporal properties of eye movements to classify visual field defects. Proceedings of the 2018 ACM Symposium on Eye Tracking Research & Applications - ETRA ’18, 2018.

Grillini, A., Renken, R. J., Vrijling, A. C. L., Heutink, J. H. C., and Cornelissen, F. W. Eye movement evaluation in multiple sclerosis and parkinson’s disease using a standardized oculomotor and neuro-ophthalmic disorder assessment (SONDA). *Preprint on medRxiv*, 2020.

Heijl, A. and Bengtsson, B. Early visual field defects in glaucoma: A study of eyes developing field loss. Glaucoma: Decision Making in Therapy, pp. 75–78, 1996.

Ishiyama, Y., Murata, H., and Asaoka, R. The usefulness of gaze tracking as an index of visual field reliability in glaucoma patients. Invest. Ophthalmol. Vis. Sci., 56(11):6233–6236, October 2015.

Jacobs, N. A. and Patterson, I. H. Variability of the hill of vision and its significance in automated perimetry. Br. J. Ophthalmol., 69(11):824–826, November 1985.

Johnson, C. A., Adams, C. W., and Lewis, R. A. Fatigue effects in automated perimetry. Applied Optics, 27(6):1030, 1988.

Jones, P. R., Smith, N. D., Bi, W., and Crabb, D. P. Portable perimetry using Eye-Tracking on a tablet Computer—A feasibility assessment. Translational Vision Science & Technology, 8(1):17, 2019.

Kim, D. E., Eizenman, M., Trope, G. E., and Kranemann, C. Eye movement perimetry. In Proceedings of 17th International Conference of the Engineering in Medicine and Biology Society, volume 2, pp. 1629–1630. IEEE, 1995.

McIlreavy, L., Fiser, J., and Bex, P. J. Impact of simulated central scotomas on visual search in natural scenes. Optom. Vis. Sci., 89(9):1385–1394, September 2012.

Montolio, F. G. J., Junoy Montolio, F. G., Wesselink, C., Gordijn, M., and Jansonius, N. M. Factors that influence standard automated perimetry test results in glaucoma: Test reliability, technician experience, time of day, and season. Investigative Opthalmology & Visual Science, 53(11):7010, 2012.

Patel, D. E., Cumberland, P. M., Walters, B. C., Russell-Eggitt, I., Rahi, J. S., and OPTIC study group. Study of optimal perimetric testing in children (OPTIC): Feasibility, reliability and repeatability of perimetry in children. PLoS One, 10(6):e0130895, June 2015.

Pel, J. J. M., van Beijsterveld, M. C. M., Thepass, G., and van der Steen, J. Validity and repeatability of saccadic response times across the visual field in eye movement perimetry. Transl. Vis. Sci. Technol., 2(7):3, November 2013.

Rottach, K. G., Riley, D. E., DiScenna, A. O., Zivotofsky, A. Z., and Leigh, R. J. Dynamic properties of horizontal and vertical eye movements in parkinsonian syndromes. Ann. Neurol., 39(3):368–377, March 1996.

Rumelhart, D. E., Hinton, G. E., and Williams, R. J. Learning representations by back-propagating errors. Nature, 323(6088):533–536, 1986.

Schultz, J. S. The “learning effect” in automated perimetry. Ophthalmology, 97(7):838, 1990.

Smith, S. and Nichols, T. Threshold-free cluster enhancement: Addressing problems of smoothing, threshold dependence and localisation in cluster inference. NeuroImage, 44(1):83–98, 2009.

Szatmáry, G. Can swedish interactive thresholding algorithm fast perimetry be used as an alternative to goldmann perimetry in neuro-ophthalmic practice? Archives of Ophthalmology, 120(9):1162, 2002.

Tieleman, T. and Hinton, G. Lecture 6.5-rmsprop: Divide the gradient by a running average of its recent magnitude. COURSERA: Neural networks for machine learning, 4(2):26–31, 2012.

Tolentino, M. J., Miller, S., Gaudio, A. R., and Sandberg, M. A. Visual field deficits in early age-related macular degeneration. Vision Res., 34(3):409–413, February 1994.

Tschopp, C., Safran, A. B., Viviani, P., Reicherts, M., Bullinger, A., and Mermoud, C. Automated visual field examination in children aged 5–8 years. part II: Normative values. Vision Res., 38(14): 2211–2218, July 1998.

Van der Stigchel, S., Bethlehem, R. A. I., Klein, B. P., Berendschot, T. T. J. M., Nijboer, T. C. W., and Dumoulin, S. O. Macular degeneration affects eye movement behavior during visual search. Front. Psychol., 4:579, September 2013.

Voipio, H. and Karjalainen, K. RETINAL AND VISUAL FIELD CHANGES IN CHLOROQUINE RETINOPATHY. Acta Ophthalmologica, 45(2):150–158, 2009.

Walters, B. C., Rahi, J. S., and Cumberland, P. M. Perimetry in children: survey of current practices in the united kingdom and ireland. Ophthalmic Epidemiol., 19(6):358–363, December 2012.

Wild, J., Kim, L., Pacey, I., and Cunliffe, I. Evidence for a learning effect in Short-Wavelength automated perimetry. Ophthalmology, 113(2):206–215, 2006.

Williams, D. Visual function in patients with homonymous hemianopia. Neurocase, 3(5):395f–403, 1997.

